# Bridging the Awareness–Utilisation Gap in Reusable Menstrual Product Use Among Female Medical Students and Healthcare Professionals: A Cross-Sectional Study

**DOI:** 10.64898/2026.04.10.26350626

**Authors:** C.F. Wami-Amadi, I.I. Nonju

**Author notes:** **Corresponding Author:** Dr Chisom Faith Wami-Amadi. **Authors’ Contributions:** C.F.W-A.: Conceptualisation, methodology, investigation, data curation, formal analysis, writing—original draft preparation and editing. I.I.N.: Investigation and validation, Writing- review All authors approved the final version of the manuscript.

## Abstract

**Background:** Reusable menstrual products provide sustainable and cost-effective alternatives to disposable sanitary products; however, their adoption remains limited, even among healthcare professionals.

**Objectives:** To assess awareness, knowledge, perceptions, and utilisation of reusable menstrual products among female medical students and healthcare professionals, and to identify predictors of willingness and use.

**Design:** Cross-sectional analytical study.

**Setting:** An online survey was conducted among female medical students and healthcare professionals in Nigeria.

**Participants:** A total of 203 female respondents aged 15–55 years.

**Intervention:** Not applicable.

**Primary Outcome Measures:** Utilisation of reusable menstrual products and willingness to adopt their use.

**Secondary Outcome Measures:** Awareness, knowledge, perceptions, and barriers.

**Methods:** Data were collected using a structured questionnaire and analysed using descriptive statistics, chi-square tests, and logistic regression.

**Results:** Awareness was high (96.06%), but utilisation was low, with 5.42% ever using and 4.43% currently using reusable products. About 31.53% were willing to use them. Respondent type was not associated with willingness (p = 0.735), although healthcare professionals had higher knowledge (p = 0.024). Positive perception predicted willingness (AOR = 7.58, 95% CI: 3.18–18.03, p < 0.001). Good knowledge (AOR = 14.96, p = 0.014) and increasing age (AOR = 1.28, p = 0.004) predicted utilisation.

**Conclusion:** Despite high awareness, utilisation remains low. Perception influences willingness, while knowledge drives use. Targeted behavioural and educational interventions are needed.

**Article Summary:** Strengths and limitations of this study

- This study used a cross-sectional design with a structured questionnaire to assess awareness, knowledge, perceptions, and utilisation of reusable menstrual products among healthcare trainees and professionals.
- The inclusion of both medical students and healthcare professionals enabled comparison across groups with differing levels of clinical exposure.
- The use of multivariable logistic regression allowed identification of independent predictors of willingness and utilisation.
- The study employed convenience sampling and an online survey, which may limit the representativeness of the sample.
- Data were self-reported, which may introduce recall and social desirability bias.

## Background

Menstrual hygiene management (MHM) is a key determinant of reproductive health and gender equity and has received increasing global attention in recent years (World Health Organisation [WHO], 2022; Hennegan et al., 2019). Reusable menstrual products, including menstrual cups, reusable sanitary pads, and menstrual discs, are designed for repeated use and offer sustainable alternatives to disposable products due to their long-term cost-effectiveness and reduced environmental impact (van Eijk et al., 2019; Kuhlmann et al., 2017).

Despite these advantages, disposable sanitary products remain the predominant choice in many settings, contributing significantly to environmental waste and recurring financial burden (Kuhlmann et al., 2017). This continued reliance reflects not only issues of accessibility and convenience but also entrenched behavioural norms and sociocultural perceptions surrounding menstruation (Hennegan et al., 2019).

Healthcare professionals and medical students occupy a unique position as both users and disseminators of reproductive health knowledge. Their personal practices and professional recommendations can significantly influence menstrual health behaviours within the broader population. However, existing evidence suggests that awareness of reusable menstrual products does not necessarily translate into their adoption, highlighting a persistent knowledge–practice gap (van Eijk et al., 2019).

Understanding this gap is particularly important in low- and middle-income contexts, where sustainable menstrual hygiene solutions could offer both economic and environmental benefits (WHO, 2022). This study, therefore, aimed to assess the utilisation, knowledge, and perceptions of reusable menstrual products among female medical students and healthcare professionals, with a view to identifying barriers and opportunities for improved adoption.

## Methods

This study employed a cross-sectional descriptive design to assess the utilisation, knowledge, and perceptions of reusable menstrual products among female medical students and healthcare professionals. The study was conducted using an online survey distributed across relevant academic and professional networks.

The study population comprised female medical students and healthcare professionals, including doctors, nurses, and allied health workers. Eligible participants were females aged 15–55 years who had experienced menstruation and provided informed consent to participate. Participants who declined consent or submitted incomplete responses were excluded from the analysis.

Participants were recruited using a convenience sampling technique through online platforms, including social media and professional communication channels. Data were collected using a structured, self-administered questionnaire developed using Kobo Toolbox Software. The questionnaire was designed based on existing literature on menstrual hygiene practices and included both closed-ended and multiple-response questions.

The instrument comprised the following sections:

Section A: Sociodemographic characteristics (age, respondent type, educational level, income, residence)

Section B: Menstrual history (cycle regularity, duration, flow characteristics, dysmenorrhea)

Section C: Awareness and knowledge of reusable menstrual products

Section D: Utilisation patterns (current and past use of menstrual products)

Section E: Willingness to adopt reusable menstrual products

Section F: Perceptions (assessed using Likert-scale items)

Section G: Barriers to use

Section H: Recommendations and educational needs

Awareness was assessed by asking participants whether they had heard of reusable menstrual products. Knowledge was evaluated using structured questions addressing safety, cost-effectiveness, environmental impact, and product longevity. Responses were categorised based on correctness and level of certainty.

Utilisation was assessed through self-reported current and past use of reusable menstrual products. Willingness to adopt these products was measured using categorical responses (yes, no, not sure).

Perceptions were assessed using Likert-scale items evaluating safety, comfort, hygiene, environmental impact, and cost-effectiveness. Barriers were identified through multiple-response questions covering informational, cultural, and practical factors.

The questionnaire was distributed electronically, and participation was voluntary. Informed consent was obtained before participation, and responses were collected anonymously to ensure confidentiality.

Data were exported into Microsoft Excel and subsequently analysed using IBM SPSS version 25. Descriptive statistics were computed and presented as frequencies and percentages for categorical variables and means with standard deviations for continuous variables. Bivariate analysis was conducted using the chi-square test and Fisher’s exact test to assess associations between categorical variables, including respondent type, knowledge level, and willingness to use reusable menstrual products.

Multivariable binary logistic regression analysis was performed to identify independent predictors of willingness to use and current utilisation of reusable menstrual products. Variables entered into the regression model included age, respondent type, knowledge level, perception, and barrier count. The primary outcomes included awareness, utilisation, and willingness to adopt reusable menstrual products. Results were presented using tables and graphical representations.

This study adhered to the STROBE reporting guidelines. (see Appendix II)

### Ethical Considerations

Ethical approval for the study was obtained from the Medical Science Ethical Committee. Participation was voluntary, and informed consent was obtained from all participants. Confidentiality and anonymity were maintained throughout the study, and no identifying information was collected.

## Results

A total of 203 respondents participated in the study, with a mean age of 23.07 ± 6.96 years. The majority were undergraduate medical students (75.37%), while healthcare professionals constituted 20.69% of the sample.

Awareness of reusable menstrual products was high (96.06%) ; however, utilisation remained low, with only 5.42% of respondents having ever used them and 4.43% currently using them.

This disparity between high awareness and low utilisation is illustrated in Figure 1. As shown in Figure 2, disposable sanitary pads remain the predominant menstrual product. Approximately 31.53% of respondents were willing to use reusable menstrual products, 36.45% were unwilling, and 28.08% were uncertain. The reported barriers to the adoption of reusable menstrual products are presented in Figure 3.

**Figure 1.**
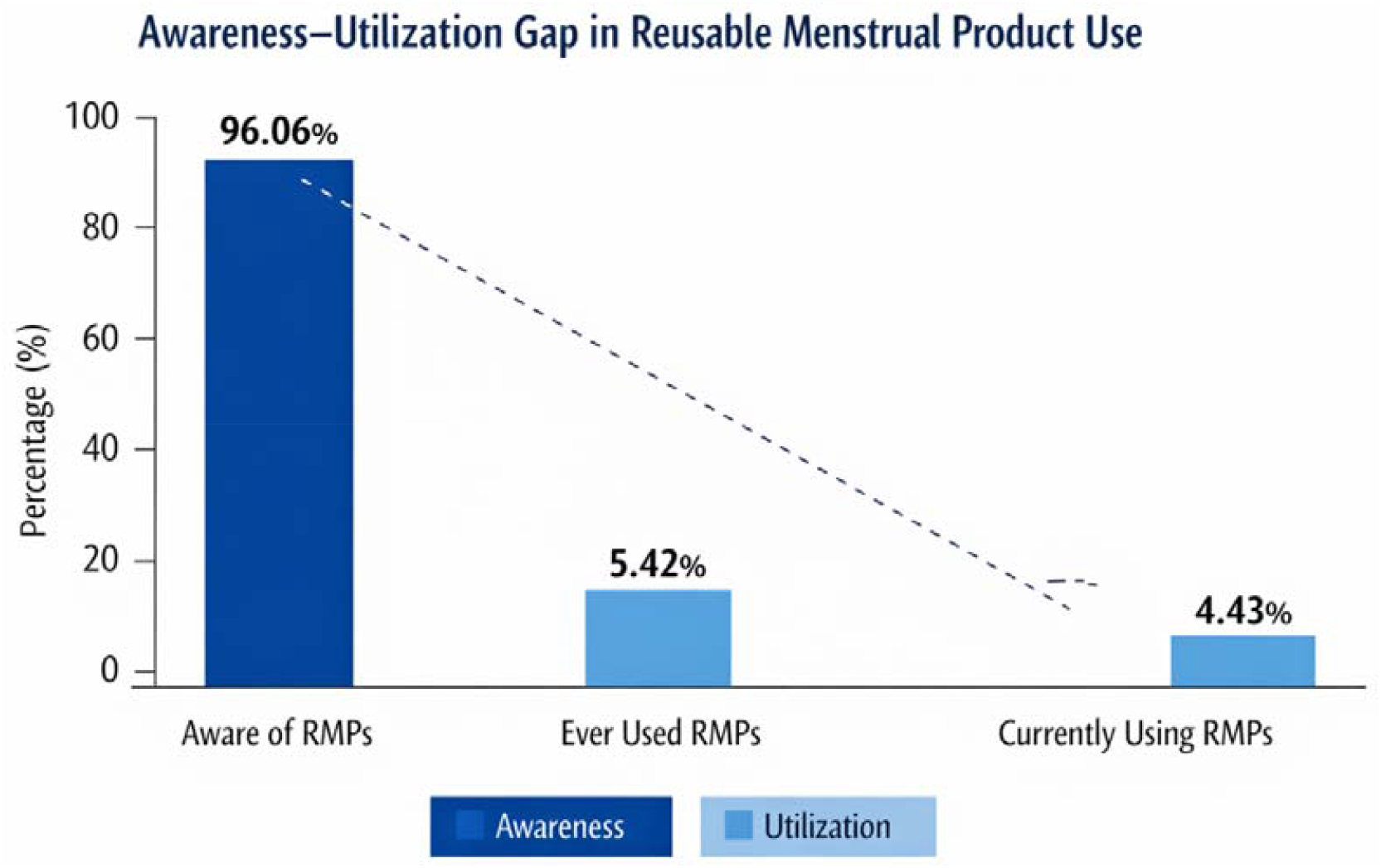
Awareness–Utilisation Gap in Reusable Menstrual Product Use Among Respondents. The figure demonstrates a substantial disparity between high awareness (96.06%) and markedly low levels of ever use (5.42%) and current use (4.43%), highlighting a critical gap between knowledge and practice.

**Figure 2.**
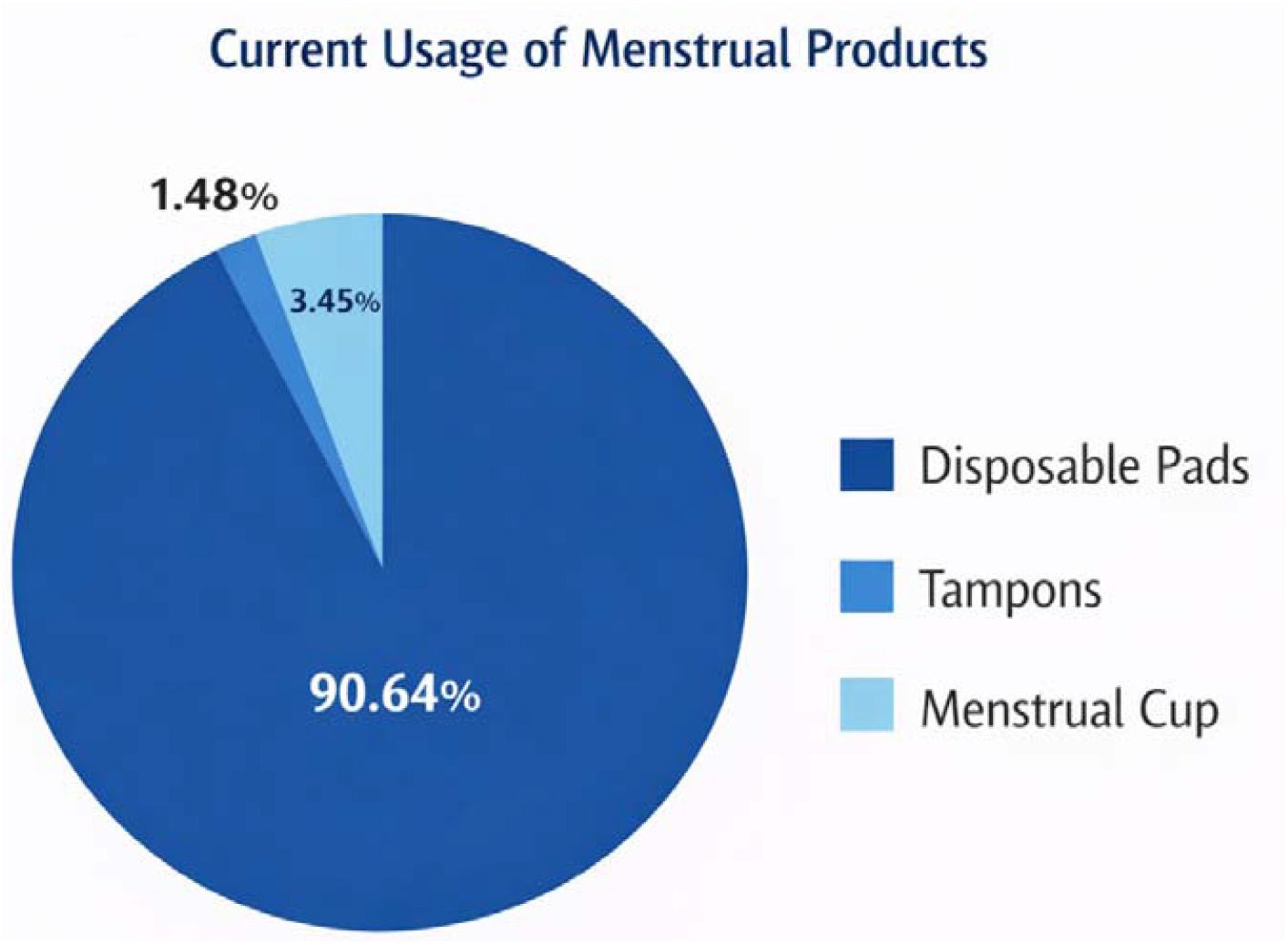
Distribution of current menstrual product use among respondents. Disposable sanitary pads were the predominant menstrual hygiene product (90.64%), with minimal use of tampons (3.45%) and menstrual cups (1.48%), reflecting low adoption of alternative menstrual products.

**Figure 3.**
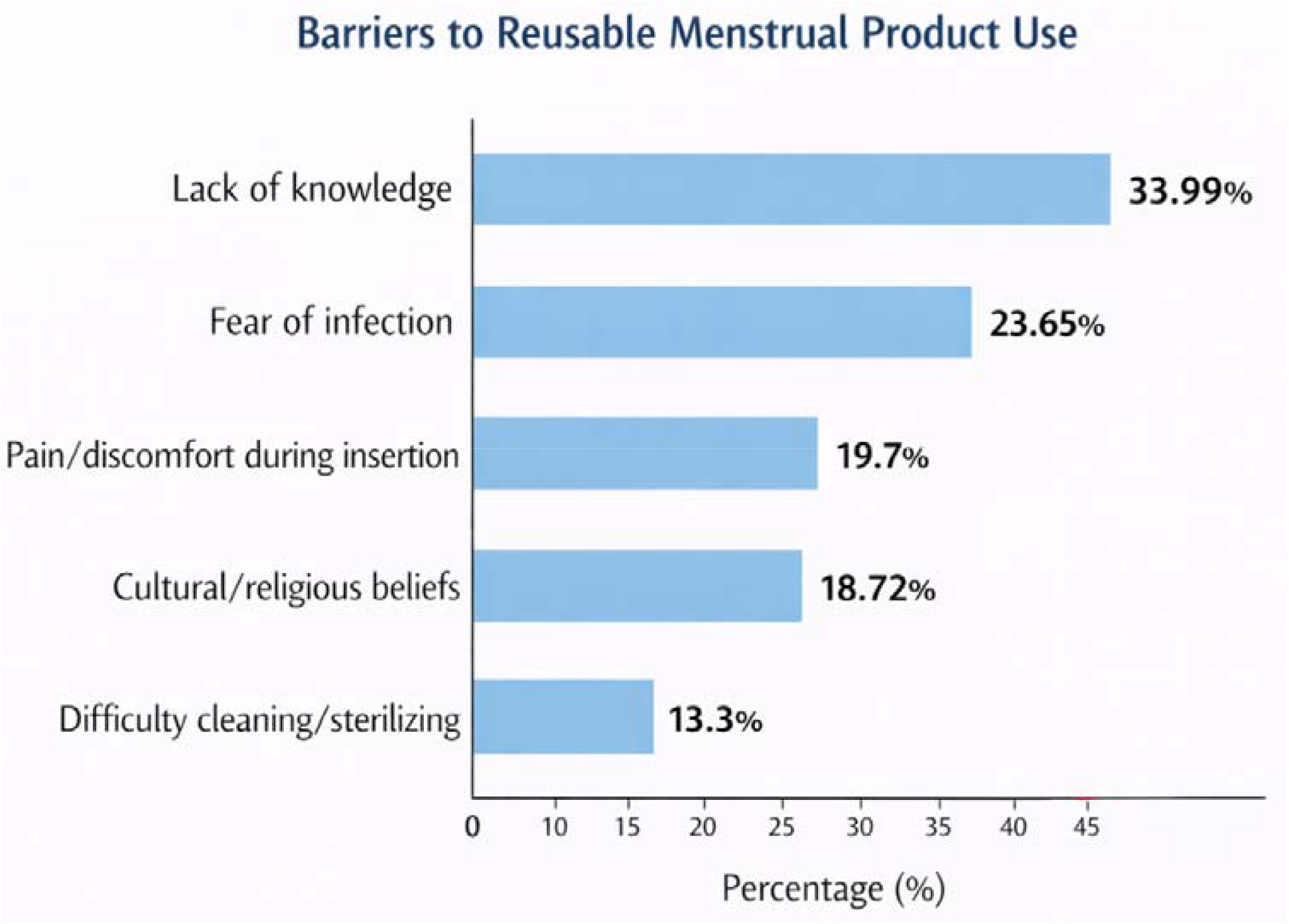
Reported Barriers to the Adoption of Reusable Menstrual Products. Key barriers included lack of knowledge, fear of infection, concerns about insertion-related discomfort, cultural or religious beliefs, and difficulty with cleaning or sterilisation, indicating both informational and sociocultural constraints.

There was no statistically significant association between respondent type (student vs healthcare professional) and willingness to use reusable menstrual products (p = 0.735). Similarly, respondent type was not significantly associated with current utilisation (p = 1.000).

However, a statistically significant association was observed between respondent type and knowledge level, with healthcare professionals more likely to demonstrate good knowledge compared to students (31.7% vs 14.8%, p = 0.024).

In multivariable logistic regression analysis, positive perception emerged as the only significant independent predictor of willingness to use reusable menstrual products (AOR = 7.58, 95% CI: 3.18–18.03, p < 0.001). Other variables, including knowledge level, respondent type, age, and barrier count, were not statistically significant predictors of willingness.

For current utilisation, good knowledge was a significant predictor (AOR = 14.96, 95% CI: 1.73–129.60, p = 0.014). Increasing age was also significantly associated with a higher likelihood of utilisation (AOR = 1.28, 95% CI: 1.08–1.51, p = 0.004).

## Discussion

These findings align with previous studies that have demonstrated the safety and acceptability of reusable menstrual products such as menstrual cups (Beksinska et al., 2015; Phillips-Howard et al., 2016). However, the present study also highlights a substantial awareness– utilisation gap. While awareness was nearly universal, actual utilisation remained remarkably low. These findings reinforce existing evidence that awareness, in isolation, is insufficient to drive meaningful behavioural change in menstrual health practices (van Eijk et al., 2019).

Importantly, inferential analysis provides deeper insight into the drivers of this gap. The lack of association between respondent type and both willingness and utilisation suggests that professional training alone does not necessarily influence menstrual product choices. Despite healthcare professionals demonstrating significantly higher knowledge levels, this did not translate into increased willingness to adopt reusable products.

One of the most notable findings of this study is that perception, rather than knowledge, was the strongest predictor of willingness to use reusable menstrual products. Participants with positive perceptions were over seven times more likely to express willingness to adopt these products. This observation is consistent with the Health Belief Model, which posits that perceived benefits and barriers influence health-related behaviours (Rosenstock, 1974).

Interestingly, while knowledge did not predict willingness, it was a significant predictor of actual utilisation. Respondents with good knowledge were substantially more likely to use reusable menstrual products. This suggests a two-step pathway: perception influences intention, while knowledge facilitates actual behaviour.

Age was also found to be a significant predictor of utilisation, with older respondents more likely to adopt reusable products. This may reflect increased autonomy, financial independence, or greater exposure to menstrual health information over time.

The absence of a significant association between healthcare professional status and utilisation is particularly noteworthy. As individuals expected to serve as health educators and advocates, this finding highlights a missed opportunity for promoting sustainable menstrual practices within clinical and educational settings.

These findings collectively suggest that interventions aimed at increasing the adoption of reusable menstrual products should not focus solely on knowledge dissemination. Instead, they should adopt a more holistic approach that addresses perceptions, cultural beliefs, and practical concerns. Strategies such as peer-led education, practical demonstrations, and healthcare provider endorsement may be particularly effective.

From a public health perspective, improving the uptake of reusable menstrual products has important implications for environmental sustainability and cost reduction. Addressing the awareness–utilisation gap among healthcare trainees and professionals could have a multiplier effect, influencing broader community practices.

However, the findings related to utilisation should be interpreted with caution due to the small number of current users, which may limit statistical power and precision of estimates.

## Conclusion

This study provides evidence that while awareness of reusable menstrual products is high among female medical students and healthcare professionals, utilisation remains significantly low. The findings highlight a critical gap between knowledge and practice, driven by a combination of informational, cultural, and practical barriers.

Addressing this gap requires more than increasing awareness; it necessitates targeted, multifaceted interventions that incorporate practical education, healthcare professional advocacy, and improved accessibility. Given the influential role of healthcare providers, empowering this group with accurate knowledge and practical experience is essential for broader societal impact.

Promoting the adoption of reusable menstrual products has important implications for sustainability, cost reduction, and improved menstrual health management. Bridging the awareness–utilisation gap is therefore not only a matter of individual behaviour change but also a public health priority.

### Limitations

This study has several limitations. The use of a cross-sectional design limits the ability to establish causality. Additionally, the reliance on convenience sampling may affect the generalizability of findings. Data were self-reported and may be subject to recall and social desirability bias. Furthermore, although inferential analysis was conducted, the small number of respondents currently using reusable menstrual products may limit the robustness of estimates related to utilisation. Future research incorporating inferential analysis and longitudinal designs is recommended.

## Acknowledgement

Department of Human Physiology, College of Medical Sciences, Rivers State University.

## Data Availability Statement

The datasets generated and/or analysed during the current study are publicly available in the Figshare repository at https://doi.org/10.6084/m9.figshare.31982160. The data are provided in anonymised form to protect participant confidentiality.

APPENDIX I

**SUPPLEMENTARY TABLE I.**
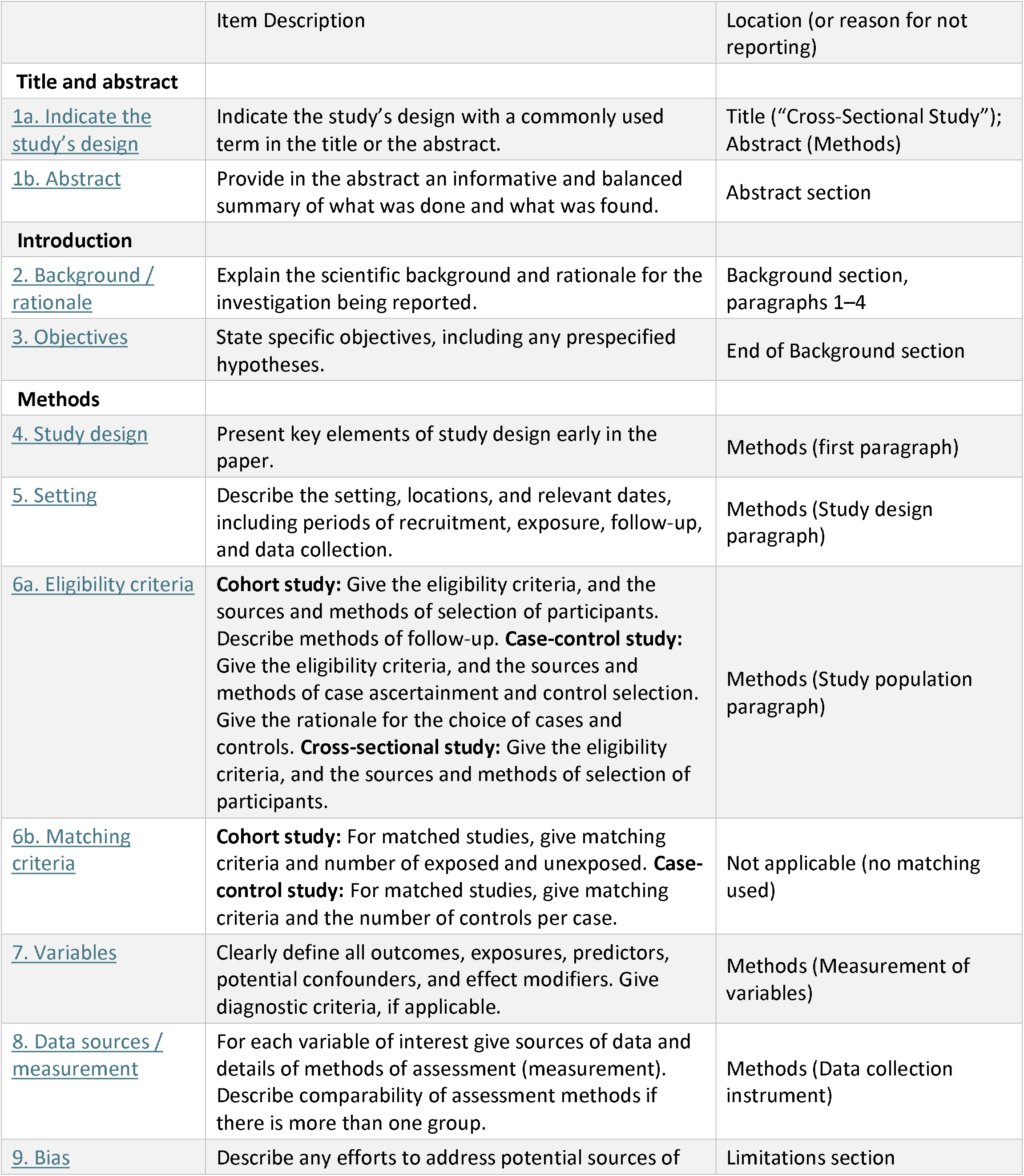
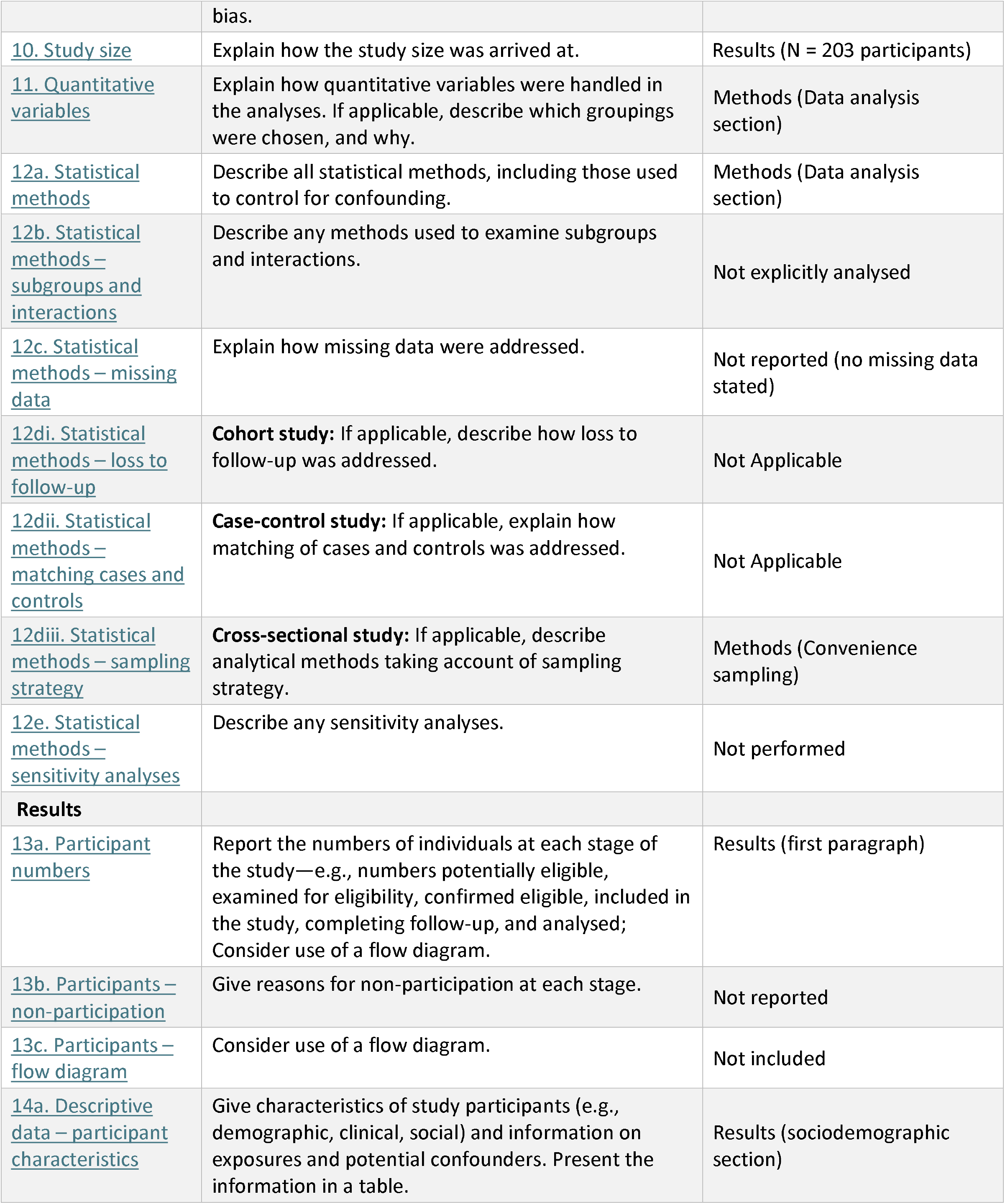
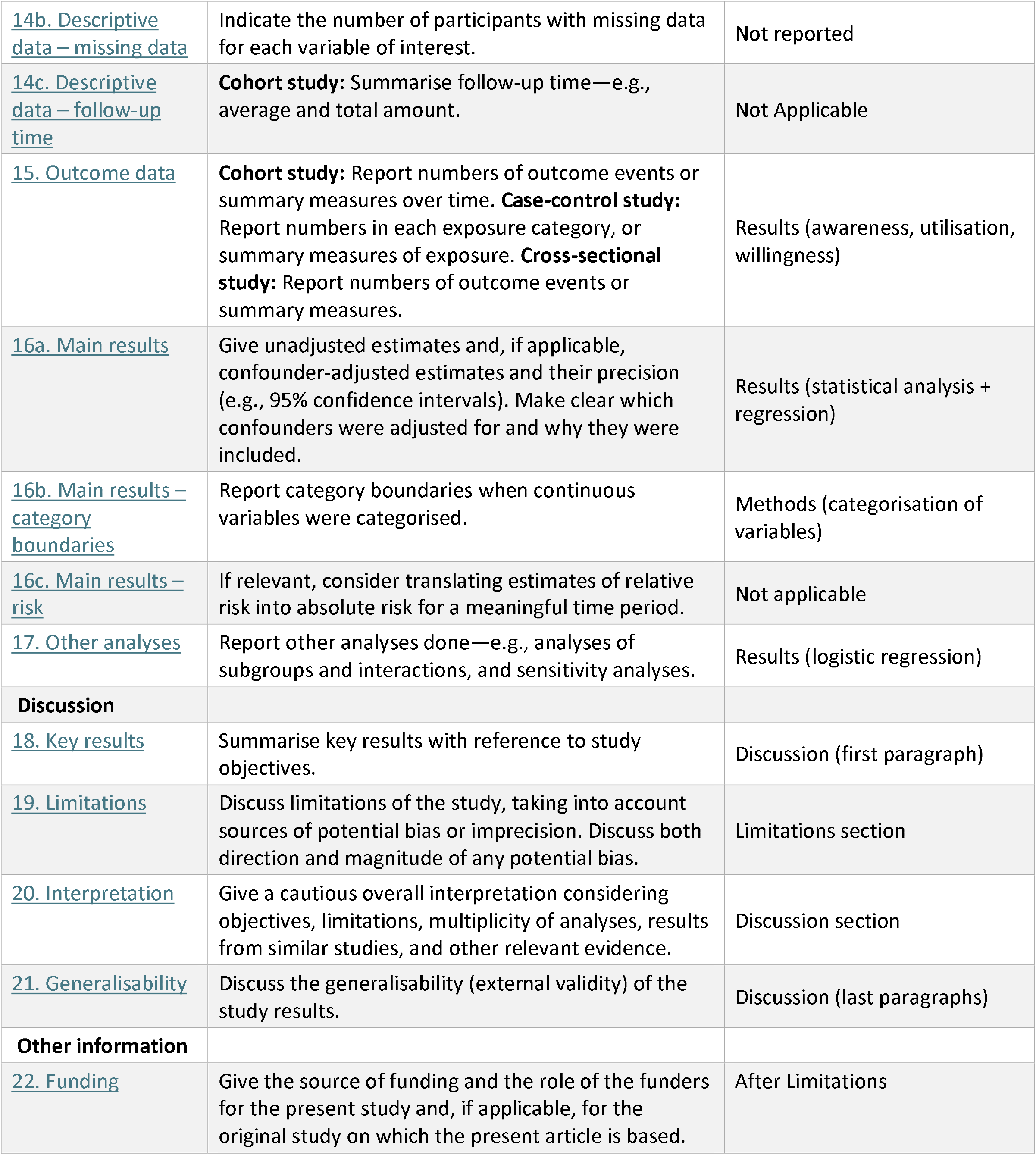
The STROBE reporting checklist.

## REFERENCES

Beksinska, M. E., Smit, J. A., & Greener, R. (2015). Acceptability and performance of the menstrual cup in South Africa: A randomised crossover trial. BMC Women’s Health, 15(1), 1–8. 10.1186/s12905-015-0183-1

Hennegan, J., Shannon, A. K., Rubli, J., Schwab, K. J., & Melendez-Torres, G. J. (2019). Women’s and girls’ experiences of menstruation in low- and middle-income countries: A systematic review and qualitative metasynthesis. PLoS Medicine, 16(5), e1002803. 10.1371/journal.pmed.1002803

Kuhlmann, A. S., Henry, K., & Wall, L. L. (2017). Menstrual hygiene management in resource-poor countries. Obstetrical & Gynaecological Survey, 72(6), 356–376. 10.1097/OGX.0000000000000443

Phillips-Howard, P. A., Nyothach, E., ter Kuile, F. O., Omoto, J., Wang, D., Zeh, C., & Mason, L. (2016). Menstrual cups and sanitary pads to reduce school attrition: A pilot study in rural Kenya. BMJ Open, 6(11), e013229. 10.1136/bmjopen-2016-013229

Rosenstock, I. M. (1974). Historical origins of the Health Belief Model. Health Education Monographs, 2(4), 328–335. 10.1177/109019817400200403

van Eijk, A. M., Zulaika, G., Lenchner, M., Mason, L., Sivakami, M., Nyothach, E., & Phillips-Howard, P. A. (2019). Menstrual cup use, leakage, acceptability, safety, and availability: A systematic review and meta-analysis. The Lancet Public Health, 4(8), e376–e393. 10.1016/S2468-2667(19)30111-2

World Health Organisation. (2022). Guidance on menstrual health and hygiene. World Health Organisation. https://www.who.int/publications/i/item/9789240043430

