## Supplemental Table 1 for "Bridging the Awareness–Utilisation Gap in Reusable Menstrual Product Use Among Female Medical Students and Healthcare Professionals: A Cross-Sectional Study"

The STROBE reporting checklist

For checking that observational epidemiology research articles can be understood and used by everyone

How to use this reporting checklist

This reporting checklist allows authors to demonstrate that their manuscripts adhere to the [STROBE reporting guideline](https:/resources.equator-network.org/reporting-guidelines/strobe/index.html).

If you have not used a reporting guideline before, read about [how and why to use them](https:/resources.equator-network.org/about/reporting-guidelines.html) and check whether STROBE is the [most applicable reporting guideline](https:/resources.equator-network.org/reporting-guidelines/strobe/index.html?#applicability) for your work.

Reporting guidelines are most useful when used early in research. When writing a manuscript or application, consider using the [full guidance](https:/resources.equator-network.org/reporting-guidelines/strobe/index.html) where you’ll find explanations and examples for each item.

After writing, demonstrate adherence by completing this checklist:

1. Specify where each item is described (see Note 1).
2. Cite this checklist (See Note 2).
3. Include your completed checklist as a supplement when submitting to a journal so that future readers can use it to find information.

|  | Item Description | Location (or reason for not reporting) |
| --- | --- | --- |
| **Title and abstract** |  |  |
| [1a. Indicate the study’s design](https:/resources.equator-network.org/reporting-guidelines/strobe/items/title-abstract-indicate-study-design.html?utm_source=strobe&utm_medium=checklist&utm_campaign=1_1) | Indicate the study’s design with a commonly used term in the title or the abstract. | Title (“Cross-Sectional Study”); Abstract (Methods) |
| [1b. Abstract](https:/resources.equator-network.org/reporting-guidelines/strobe/items/abstract.html?utm_source=strobe&utm_medium=checklist&utm_campaign=1_1) | Provide in the abstract an informative and balanced summary of what was done and what was found. | Abstract section |
| **Introduction** |  |  |
| [2. Background / rationale](https:/resources.equator-network.org/reporting-guidelines/strobe/items/background-rationale.html?utm_source=strobe&utm_medium=checklist&utm_campaign=1_1) | Explain the scientific background and rationale for the investigation being reported. | Background section, paragraphs 1–4 |
| [3. Objectives](https:/resources.equator-network.org/reporting-guidelines/strobe/items/objectives.html?utm_source=strobe&utm_medium=checklist&utm_campaign=1_1) | State specific objectives, including any prespecified hypotheses. | End of Background section |
| **Methods** |  |  |
| [4. Study design](https:/resources.equator-network.org/reporting-guidelines/strobe/items/study-design.html?utm_source=strobe&utm_medium=checklist&utm_campaign=1_1) | Present key elements of study design early in the paper. | Methods (first paragraph) |
| [5. Setting](https:/resources.equator-network.org/reporting-guidelines/strobe/items/setting.html?utm_source=strobe&utm_medium=checklist&utm_campaign=1_1) | Describe the setting, locations, and relevant dates, including periods of recruitment, exposure, follow-up, and data collection. | Methods (Study design paragraph) |
| [6a. Eligibility criteria](https:/resources.equator-network.org/reporting-guidelines/strobe/items/eligibility-criteria.html?utm_source=strobe&utm_medium=checklist&utm_campaign=1_1) | **Cohort study:** Give the eligibility criteria, and the sources and methods of selection of participants. Describe methods of follow-up. **Case-control study:** Give the eligibility criteria, and the sources and methods of case ascertainment and control selection. Give the rationale for the choice of cases and controls. **Cross-sectional study:** Give the eligibility criteria, and the sources and methods of selection of participants. | Methods (Study population paragraph) |
| [6b. Matching criteria](https:/resources.equator-network.org/reporting-guidelines/strobe/items/matching-criteria.html?utm_source=strobe&utm_medium=checklist&utm_campaign=1_1) | **Cohort study:** For matched studies, give matching criteria and number of exposed and unexposed. **Case-control study:** For matched studies, give matching criteria and the number of controls per case. | Not applicable (no matching used) |
| [7. Variables](https:/resources.equator-network.org/reporting-guidelines/strobe/items/variables.html?utm_source=strobe&utm_medium=checklist&utm_campaign=1_1) | Clearly define all outcomes, exposures, predictors, potential confounders, and effect modifiers. Give diagnostic criteria, if applicable. | Methods (Measurement of variables) |
| [8. Data sources / measurement](https:/resources.equator-network.org/reporting-guidelines/strobe/items/data-sources-measurement.html?utm_source=strobe&utm_medium=checklist&utm_campaign=1_1) | For each variable of interest give sources of data and details of methods of assessment (measurement). Describe comparability of assessment methods if there is more than one group. | Methods (Data collection instrument) |
| [9. Bias](https:/resources.equator-network.org/reporting-guidelines/strobe/items/bias.html?utm_source=strobe&utm_medium=checklist&utm_campaign=1_1) | Describe any efforts to address potential sources of bias. | Limitations section |
| [10. Study size](https:/resources.equator-network.org/reporting-guidelines/strobe/items/study-size.html?utm_source=strobe&utm_medium=checklist&utm_campaign=1_1) | Explain how the study size was arrived at. | Results (N = 203 participants) |
| [11. Quantitative variables](https:/resources.equator-network.org/reporting-guidelines/strobe/items/quantitative-variables.html?utm_source=strobe&utm_medium=checklist&utm_campaign=1_1) | Explain how quantitative variables were handled in the analyses. If applicable, describe which groupings were chosen, and why. | Methods (Data analysis section) |
| [12a. Statistical methods](https:/resources.equator-network.org/reporting-guidelines/strobe/items/statistical-methods-description.html?utm_source=strobe&utm_medium=checklist&utm_campaign=1_1) | Describe all statistical methods, including those used to control for confounding. | Methods (Data analysis section) |
| [12b. Statistical methods – subgroups and interactions](https:/resources.equator-network.org/reporting-guidelines/strobe/items/statistical-methods-subgroups-interactions.html?utm_source=strobe&utm_medium=checklist&utm_campaign=1_1) | Describe any methods used to examine subgroups and interactions. | Not explicitly analysed |
| [12c. Statistical methods – missing data](https:/resources.equator-network.org/reporting-guidelines/strobe/items/statistical-methods-missing-data.html?utm_source=strobe&utm_medium=checklist&utm_campaign=1_1) | Explain how missing data were addressed. | Not reported (no missing data stated) |
| [12di. Statistical methods – loss to follow-up](https:/resources.equator-network.org/reporting-guidelines/strobe/items/statistical-methods-loss-to-follow-up.html?utm_source=strobe&utm_medium=checklist&utm_campaign=1_1) | **Cohort study:** If applicable, describe how loss to follow-up was addressed. | Not Applicable |
| [12dii. Statistical methods – matching cases and controls](https:/resources.equator-network.org/reporting-guidelines/strobe/items/statistical-methods-matching-cases-controls.html?utm_source=strobe&utm_medium=checklist&utm_campaign=1_1) | **Case-control study:** If applicable, explain how matching of cases and controls was addressed. | Not Applicable |
| [12diii. Statistical methods – sampling strategy](https:/resources.equator-network.org/reporting-guidelines/strobe/items/statistical-methods-analytical-methods-sampling-strategy.html?utm_source=strobe&utm_medium=checklist&utm_campaign=1_1) | **Cross-sectional study:** If applicable, describe analytical methods taking account of sampling strategy. | Methods (Convenience sampling) |
| [12e. Statistical methods – sensitivity analyses](https:/resources.equator-network.org/reporting-guidelines/strobe/items/statistical-methods-sensitivity-analyses.html?utm_source=strobe&utm_medium=checklist&utm_campaign=1_1) | Describe any sensitivity analyses. | Not performed |
| **Results** |  |  |
| [13a. Participant numbers](https:/resources.equator-network.org/reporting-guidelines/strobe/items/participants-numbers.html?utm_source=strobe&utm_medium=checklist&utm_campaign=1_1) | Report the numbers of individuals at each stage of the study—e.g., numbers potentially eligible, examined for eligibility, confirmed eligible, included in the study, completing follow-up, and analysed; Consider use of a flow diagram. | Results (first paragraph) |
| [13b. Participants – non-participation](https:/resources.equator-network.org/reporting-guidelines/strobe/items/participants-non-participation.html?utm_source=strobe&utm_medium=checklist&utm_campaign=1_1) | Give reasons for non-participation at each stage. | Not reported |
| [13c. Participants – flow diagram](https:/resources.equator-network.org/reporting-guidelines/strobe/items/participants-flow-diagram.html?utm_source=strobe&utm_medium=checklist&utm_campaign=1_1) | Consider use of a flow diagram. | Not included |
| [14a. Descriptive data – participant characteristics](https:/resources.equator-network.org/reporting-guidelines/strobe/items/descriptive-data-participant-characteristics.html?utm_source=strobe&utm_medium=checklist&utm_campaign=1_1) | Give characteristics of study participants (e.g., demographic, clinical, social) and information on exposures and potential confounders. Present the information in a table. | Results (sociodemographic section) |
| [14b. Descriptive data – missing data](https:/resources.equator-network.org/reporting-guidelines/strobe/items/descriptive-data-missing-data.html?utm_source=strobe&utm_medium=checklist&utm_campaign=1_1) | Indicate the number of participants with missing data for each variable of interest. | Not reported |
| [14c. Descriptive data – follow-up time](https:/resources.equator-network.org/reporting-guidelines/strobe/items/descriptive-data-follow-up-time.html?utm_source=strobe&utm_medium=checklist&utm_campaign=1_1) | **Cohort study:** Summarise follow-up time—e.g., average and total amount. | Not Applicable |
| [15. Outcome data](https:/resources.equator-network.org/reporting-guidelines/strobe/items/outcome-data.html?utm_source=strobe&utm_medium=checklist&utm_campaign=1_1) | **Cohort study:** Report numbers of outcome events or summary measures over time. **Case-control study:** Report numbers in each exposure category, or summary measures of exposure. **Cross-sectional study:** Report numbers of outcome events or summary measures. | Results (awareness, utilisation, willingness) |
| [16a. Main results](https:/resources.equator-network.org/reporting-guidelines/strobe/items/main-results.html?utm_source=strobe&utm_medium=checklist&utm_campaign=1_1) | Give unadjusted estimates and, if applicable, confounder-adjusted estimates and their precision (e.g., 95% confidence intervals). Make clear which confounders were adjusted for and why they were included. | Results (statistical analysis + regression) |
| [16b. Main results – category boundaries](https:/resources.equator-network.org/reporting-guidelines/strobe/items/main-results-category-boundaries.html?utm_source=strobe&utm_medium=checklist&utm_campaign=1_1) | Report category boundaries when continuous variables were categorised. | Methods (categorisation of variables) |
| [16c. Main results – risk](https:/resources.equator-network.org/reporting-guidelines/strobe/items/main-results-risk.html?utm_source=strobe&utm_medium=checklist&utm_campaign=1_1) | If relevant, consider translating estimates of relative risk into absolute risk for a meaningful time period. | Not applicable |
| [17. Other analyses](https:/resources.equator-network.org/reporting-guidelines/strobe/items/other-analyses.html?utm_source=strobe&utm_medium=checklist&utm_campaign=1_1) | Report other analyses done—e.g., analyses of subgroups and interactions, and sensitivity analyses. | Results (logistic regression) |
| **Discussion** |  |  |
| [18. Key results](https:/resources.equator-network.org/reporting-guidelines/strobe/items/key-results.html?utm_source=strobe&utm_medium=checklist&utm_campaign=1_1) | Summarise key results with reference to study objectives. | Discussion (first paragraph) |
| [19. Limitations](https:/resources.equator-network.org/reporting-guidelines/strobe/items/limitations.html?utm_source=strobe&utm_medium=checklist&utm_campaign=1_1) | Discuss limitations of the study, taking into account sources of potential bias or imprecision. Discuss both direction and magnitude of any potential bias. | Limitations section |
| [20. Interpretation](https:/resources.equator-network.org/reporting-guidelines/strobe/items/interpretation.html?utm_source=strobe&utm_medium=checklist&utm_campaign=1_1) | Give a cautious overall interpretation considering objectives, limitations, multiplicity of analyses, results from similar studies, and other relevant evidence. | Discussion section |
| [21. Generalisability](https:/resources.equator-network.org/reporting-guidelines/strobe/items/generalisability.html?utm_source=strobe&utm_medium=checklist&utm_campaign=1_1) | Discuss the generalisability (external validity) of the study results. | Discussion (last paragraphs) |
| **Other information** |  |  |
| [22. Funding](https:/resources.equator-network.org/reporting-guidelines/strobe/items/funding.html?utm_source=strobe&utm_medium=checklist&utm_campaign=1_1) | Give the source of funding and the role of the funders for the present study and, if applicable, for the original study on which the present article is based. | After Limitations |

### 1 How to specify where content is

Tell the reader where they can find information. E.g.,

- Results; paragraph 2
- Methods, Participants; paragraphs 1 & 2.
- Table 3
- Supplement B, para. 4

If you have chosen not to describe an item, explain why. You can do this in the checklist, or as a note below it.

You can describe items in the article body, or in tables, figures, or supplementary materials, and should prioritize items you feel are most important to your intended audience. The order of items in your manuscript does not need to match the order of items in this checklist. You can decide how best to structure your work.

### 2 How to cite

Describe how you used STROBE at the end of your Methods section, referencing the resources you used e.g.,

‘We used the STROBE reporting guideline(1) to draft this manuscript, and the STROBE reporting checklist(2) when editing, included in supplement A’

If you use a reporting checklist, remember to include it as a supplement when publishing so that readers can easily find information and see how you have interpreted the guidance.

1.	Elm E von, Altman DG, Egger M, Pocock SJ, Gøtzsche PC, Vandenbroucke JP, et al. The strengthening the reporting of observational studies in epidemiology (STROBE) statement: Guidelines for reporting observational studies. Annals of Internal Medicine [Internet]. 2007 Oct;147(8):573–7. Available from: <https://www.acpjournals.org/doi/10.7326/0003-4819-147-8-200710160-00010>

2.	Elm E von, Altman DG, Egger M, Pocock SJ, Gøtzsche PC, Vandenbroucke JP, et al. The STROBE reporting checklist. In: Harwood J, Albury C, Beyer J de, Schlüssel M, Collins G, editors. The EQUATOR network reporting guideline platform [Internet]. The UK EQUATOR Centre; 2025. Available from: [https:/resources.equator-network.org/reporting-guidelines/strobe/strobe-checklist.docx](https://https:/resources.equator-network.org/reporting-guidelines/strobe/strobe-checklist.docx)
